# Patient’s Satisfaction with Eye Care Services from tertiary Eye hospital in Nepal

**DOI:** 10.1101/2023.10.18.23297243

**Authors:** Ranjan Shah, Sailesh Kumar Mishra, Prakriti Sharma

**Affiliations:** Nepal Netra Jyoti Sangh, Kathmandu (NNJS), Nepal; Nobel College, Sinamangal

**Keywords:** Patient’s satisfaction, Eye Care Service, Tertiary eye hospital, Madhesh Province

## Abstract

**Purpose:** This study was conducted to assess the level of patient satisfaction with the eye care services received from different tertiary eye hospitals, running under Nepal Nepal Netra Jyoti (NNJS) Sangh, of Madhesh Province, Nepal over a 3-month period (October-December) 2017.

**Methods:** A Cross-sectional based study with client exit interviews were done, using pretested Likert scale tool in sample of 1190 (P=0.5, and 95% CI) consenting patients after receiving the ophthalmic care enrolled in the mentioned tertiary eye hospitals of Madhesh Province. Data on Behavior of the service provider, Services and facilities provided by the hospital was analyzed with the level of patient’s satisfaction. A Pearson’s chi-square test was used to examine the relationship between overall satisfaction and the dependent variables.

**Results:** Only 3.8 % of patients reported visiting the hospital more than four times. The main reasons for choosing their respective hospital was the ‘quality of services’ delivered through modern equipment at affordable costs (53 %) followed by easy access to hospital (39%). The average waiting time was 58 min, with a range of 5-240 min. Level of education (P<0.05), Socioeconomic status (P<0.05), Waiting time (P<0.05), Hospital operating hours (P<0.05), Cleanliness (P<0.05) and Cost of Service (P<0.05) were significantly associated with the overall satisfaction with the services. Behavior of the service providers did not play a statistically significant role in overall satisfaction.

**Conclusion:** Patient satisfaction is a highly desirable outcome of clinical care and may even be an element of health status itself. Our data showed that a significant proportion (95%) of patients were satisfied, or highly satisfied overall (across all parametric measures), and that personal experience, beliefs and background may play a confounding factor in satisfaction. As a result, it can be safely concluded that perceived quality and performance in the hospitals are exceptional.

## Introduction

Patient satisfaction refers to how closely a patient’s expectations of the optimum level of care match up with how they feel they actually received that level of care^1^. In recent years, patient happiness has gained in prominence. Many healthcare professionals are starting to recognize its use in evaluating hospital performance, quality, and patient outcomes. These fundamental factors are worldwide problems that several nations at all stages of. As a result, the health care sector is rapidly changing to meet the rising demands and needs of its patient population^2,3^. Standards of professional practice and other provider/hospital-oriented indicators, such as technology, have historically been used to measure the quality of healthcare. However, there is less of a focus on objective statistics and more on subjective empirical evidence as a result of this renewed interest in patient-oriented indicators. Moreover, health care recipients in developing countries are more sensitive to perceptions of the quality of healthcare service^4^.

Out of 77 districts of Nepal, 60 of them provides eye care services under NNJS where the users exceed 3 million every year. The number of eye care professionals has expanded to 420 ophthalmologists, over 2000 ophthalmic assistants, and more than 1300 optometrists, with a cataract surgical rate of 4500. As per the Rapid Assessment of avoidable blindness survey in 2021, the National CSC_**<6/60**_ for the total population 50 years and above is 82.7% (95% CI 80.8 - 84.4) which is slightly less than the target of at least 85% recommended by the International Agency for the Prevention of Blindness (IAPB).

Patient satisfaction surveys examine many and novel facets of healthcare, offering a wonderful chance to involve clients or patients in the process of enhancing the standard of services and, consequently, the quality of patients’ lives^5^. Previous studies suggest improvement in communication skills, technical help as well as an alternative option for funding to the patient without health insurance for patient’s satisfaction^6^.

Despite the prevalence of patient satisfaction studies in ophthalmology care around the world, relatively few studies have been undertaken in Nepal. In contrast, many patient satisfaction studies were conducted in Nepal to evaluate health services in other fields of medicine but in regards to eye services very few studies have been conducted. Thus this study aims to analyse the level of patient satisfaction with eye care services received from tertiary hospital of Madhesh Province

## Materials and methods

### Study design and settings

A cross-sectional descriptive study conducted in three different eye hospitals of Madhesh Province which are running under NNJS. The hospitals were selected from 3 different districts (Rautahat, Bara and Siraha). They were Gaur eye hospital, R. M. Kedia eye hospital, Sagarmatha Choudhary eye Hospital respectively. The patient getting Ophthalmic care services from OPD and IPD (Out-patient and in-patient department) were enrolled in this study.

### Ethics and consent

Ethical approval for the study was provided by ethical review board committee of Nepal health Research Council (NHRC). Permission was granted by the NNJS central office which run these eye hospitals prior to the study. Participant were provided a detailed explanation of the purpose and scope along with formal and written consent before including them in this study. Before the data collection, written consent was taken from each participants. All the participants were informed about their right to withdraw from the research if they are not comfortable providing the information. In case of under 18 years of age participants, their parents and guardians were interviewed and identity of the participation was kept confidential.

### Study procedure

The study was carried out between October and December 2017. The previous studies on patient satisfaction with eye care service and using appropriate statistical formula for estimating sample size [n=Z pq/d] with 5% non-responsive rate, a sample of 403 per hospital (1209 from the three hospital) was calculated to detect level of satisfaction among the study participant. Structured questionnaire was used with 5 point Likert scale ranging from Highly dissatisfied to Highly satisfied. Open and close ended questionnaire was used developed in English and later translated in Nepali for convenience. A pilot test was conducted in 50 patients in Kritipur Hospital for validity which was not included in this study. The questionnaire consists of 34 items which measures socio-demographic variable, several core dimension of patient satisfaction, behaviour of service provider, Services and facilities provided in OPD and IPD department.

### Data analysis

The data was cleaned, edited and coded in the same day of data collection. Data entry and analysis was carried out in SPSS (Statistical Package for the Social Sciences). Descriptive Analysis was done to check distribution of data, detecting outliers and measure association among variables. Chi-square test was done in Bivariate analysis to identify association between dependent and independent variables.

## Results and discussion

### Descriptive analysis

Among 1190 respondents, the average male and female population in selected tertiary hospitals were 66% and 33% respectively. The mean age was 40 years and 79% of patients were from out-patient-department and 21% from the inpatient department. Among the three hospitals, Gaur Hospital had the highest proportion of neutrality or dissatisfaction in compared to other hospitals. Table 1 shows the overall satisfaction in aggregate of 3 different hospitals’, demographic characteristics of the patient included in the study. In regards to the educational qualification, majority of the population were illiterate or could only read and write whereas for survival length based on respondent’s main source of income 67.06% reported less than 6 months. Very less respondents had visited 4 times or more for eye check up in the hospital and their major reason for hospital visit was for quality of service by modern equipment at low price.

**Table 1:** Socio-demographic Variable.

| Variable | Negative | Positive | Total |
| --- | --- | --- | --- |
| **Sex** |  |  |  |
| Male | 37 | 757 | 794 |
| Female | 22 | 374 | 396 |
| **Age** |  |  |  |
| Under 18 | - | 7 | 7 |
| 19 -29 | 2 | 88 | 90 |
| 30-39 | 13 | 204 | 217 |
| 40-49 | 17 | 366 | 383 |
| 50-59 | 9 | 243 | 252 |
| 60-69 | 16 | 206 | 222 |
| 70-79 | - | 25 | 25 |
| 80-89 | - | 5 | 5 |
| 90-99 | - | 4 | 4 |
| **Patient type** |  |  |  |
| Out patient | 43 | 901 | 944 |
| inpatient | 16 | 230 | 246 |
| **Education level** |  |  |  |
| Illiterate | 24 | 365 | 388 |
| only read and write | 26 | 354 | 380 |
| primary level education | 5 | 205 | 210 |
| lower secondary level | 3 | 99 | 102 |
| secondary level | 1 | 89 | 90 |
| higher education or more | - | 20 | 20 |
| **Length of survival on main source of income** |  |  |  |
| Less than 6 months | 54 | 744 | 798 |
| 6-12 months | 4 | 190 | 194 |
| More than 1 year with saving | 1 | 197 | 198 |
| **Number of visits** |  |  |  |
| 1<sup>st</sup> time | 31 | 556 | 587 |
| 2<sup>nd</sup> time | 22 | 460 | 482 |
| 3<sup>rd</sup> time | 4 | 71 | 75 |
| 4<sup>th</sup> or more | 2 | 44 | 46 |
| **Reason for check-up** |  |  |  |
| Qualified doctors, nurses, examiners and staff | 2 | 71 | 73 |
| Quality services by modern equipment for low price | 25 | 610 | 635 |
| Attractive hospital building | 1 | 13 | 14 |
| Easy access | 31 | 437 | 468 |

\*Positive response only includes those who responded “SFD” or “HSFD” to question of overall satisfaction

In regards to the behaviour of service provider and service and facility related variable, majority of respondents expressed satisfaction with the behaviour of the doctor’s/examiners, registration personnel and spectacle dispensers. Moreover, at least 80% of patients reported being SFD or HSFD (satisfied or highly satisfied), thus service- and facility-related characteristics were also highly appreciated. In all three selected hospitals, high number of patients were satisfied with the services provided by the hospital. Table 2 and table 3 shows the level of satisfaction among the three hospital.

**Table 2:** Behaviour of service provider.

| Variable | Negative | Positive | Total |
| --- | --- | --- | --- |
| **Behavior of the staff at the registration department of the hospital** |  |  |  |
| Highly dissatisfied | - | 1 | 1 |
| Dissatisfied | - | - | - |
| Fairly | 2 | 21 | 23 |
| Satisfied | 33 | 694 | 727 |
| Highly satisfied | 24 | 415 | 493 |
| **Behavior of doctors and examiners** |  |  |  |
| Fairly | 54 | 744 | 798 |
| Satisfied | 4 | 190 | 194 |
| Highly satisfied | 1 | 197 | 198 |
| **Behavior of staff in pharmaceutical and spectacle dispensing unit** |  |  |  |
| Highly dissatisfied | 31 | 556 | 587 |
| Dissatisfied | 22 | 460 | 482 |
| Fairly | 4 | 71 | 75 |
| Satisfied | 2 | 44 | 46 |

\*Positive response only includes those who responded “SFD” or “HSFD” to question of overall satisfaction

**Table 3:** Service and Facilitators.

| Variable | Negative | Positive | Total |
| --- | --- | --- | --- |
| **Opening hours/days of hospital** |  |  |  |
| Highly dissatisfied | - | - | - |
| Dissatisfied | 3 | 7 | 10 |
| Fairly | 2 | 63 | 65 |
| Satisfied | 41 | 699 | 740 |
| Highly satisfied | 13 | 362 | 375 |
| **Waiting time** |  |  |  |
| 1190 Observations | Mean (57.5) | Std Dev (29.2) | Minimum (5)/<br>Maximum (240) |
| **Attention given by examiners** |  |  |  |
| Highly dissatisfied | - | - | - |
| Dissatisfied | 1 | 2 | 3 |
| Fairly | 2 | 12 | 14 |
| Satisfied | 23 | 508 | 531 |
| Highly satisfied | 33 | 609 | 642 |
| **Examination and treatment suggestion** |  |  |  |
| Highly dissatisfied | - | - | - |
| Dissatisfied | 1 | 1 | 2 |
| Fairly | 1 | 29 | 30 |
| Satisfied | 24 | 576 | 600 |
| Highly satisfied | 33 | 525 | 558 |
| **Cleanliness of OPD** |  |  |  |
| Highly dissatisfied | - | - | - |
| Dissatisfied | 1 | 5 | 6 |
| Fairly | 9 | 152 | 161 |
| Satisfied | 42 | 655 | 697 |
| Highly satisfied | 7 | 319 | 326 |
| **Pharmaceutical and spectacle dispensing unit** |  |  |  |
| Highly dissatisfied | - | - | - |
| Dissatisfied | - | 2 | 2 |
| Fairly | 15 | 146 | 161 |
| Satisfied | 37 | 658 | 695 |
| Highly satisfied | 7 | 325 | 332 |
| **Cost of service** |  |  |  |
| Highly dissatisfied | - | - | - |
| Dissatisfied | 3 | 3 | 6 |
| Fairly | 4 | 29 | 33 |
| Satisfied | 38 | 767 | 805 |
| Highly satisfied | 14 | 332 | 346 |
| **Facilities for visitors** |  |  |  |
| Highly dissatisfied | - | - | - |
| Dissatisfied | 1 | 5 | 6 |
| Fairly | 1 | 37 | 38 |
| Satisfied | 14 | 153 | 167 |
| Highly satisfied | - | 37 | 37 |
| **Cleanliness of IPD** |  |  |  |
| Highly dissatisfied | - | - | - |
| Dissatisfied | - | - | - |
| Fairly | 2 | 26 | 28 |
| Satisfied | 14 | 149 | 163 |
| Highly satisfied | 1 | 56 | 57 |
| **Behavior of nurses** |  |  |  |
| Highly dissatisfied | - | 1 | 1 |
| Dissatisfied | - | - | - |
| Fairly | 6 | 29 | 35 |
| Satisfied | 10 | 153 | 163 |
| Highly satisfied | - | 49 | 49 |
| **Performance of physicians**<br>**who conducted surgery** |  |  |  |
| Highly dissatisfied | 2 | - | 2 |
| Dissatisfied | 7 | 28 | 35 |
| Fairly | 1 | 32 | 33 |
| Satisfied | 5 | 86 | 92 |
| Highly satisfied | 1 | 85 | 86 |
| **Would you recommend this**<br>**hospital to others?** |  |  |  |
| yes | 58 | 1,131 | 1,189 |
| no | 1 | - | 1 |

\*Positive response only includes those who responded "SFD" or "HSFD" to question of overall satisfaction

### Bivariate Analysis

Majority of the respondents in selected hospital responded the service provided are fair, satisfied or highly satisfied. Pearson’s Chi-Square test was done to determine the significance level of satisfaction among the respondents. The study was significantly associated with the overall satisfaction (p= 0.004). Another factor that was significantly associated with overall satisfaction was socioeconomic level (p=.000). In comparison to patients who could survive longer than one year, those who reported having less than six months left on their primary source of income were 14% more likely to feel FR, SFD, or HSFD with the institution as a whole. In regards to behaviour of the service provider, the patient’s satisfaction was statistically insignificant but factors related to the service itself and the physical infrastructure were statistically significant. Waiting time (p=0.005), hospital working hours (p=0.001), impression of treatment plan (p=0.012), cleanliness (p=.029), and cost of service (p=.000) were all substantially correlated with level of satisfaction.

## Discussion

According to the study’s findings, patient satisfaction was generally quite high. Across all parametric metrics, covering the three institutions, 95% of patients said they were “SFD” or “HSFD” with their overall experience. The consequence is that patients believe the perceived quality to be excellent.

When decisions are made concerning service adjustments and improvements, patient satisfaction is vital, hence enhancing patient’s satisfaction may help to increase the chances that they can suggests a certain provider to friends and family who are looking for a qualified physician. For the most part, individuals trust their friends and family for information, especially when choosing a healthcare practitioner^7,8^. Roughly 99% of respondents in our study stated they would refer the hospital in question to others. Although patient happiness and referrals are connected metrics, some researchers have discovered that a hospital with high satisfaction does not always translate into high referrals, and the opposite is also true. On the other hand, only one survey participant said they would not recommend the facility to others, even though 5% of respondents (n=59) said they were generally “DSFD” or “HDSFD.” This indicates that evaluations of patient satisfaction and recommendation has to be conducted independently^9^. The majority of this report’s focus was on patient satisfaction with regard to quality and the variables influencing it.

The Kano model, developed by researchers in University of Tokyo is useful to analyse patient’s satisfaction, quality visually and allows for studying different levels of patients expectation^10^. The model demonstrates three relationships between satisfaction and quality (expected, performer and delighter). Patients will have a foundational set of unspoken expectations regarding their care that are frequently taken for granted (red line). Since they are routine, these expectations are constant. The majority of patients don’t even perceive them as comparable quality variables, thus their absence will surprise them^11^. Although the patient may not be aware of these expectations, if they are not fulfilled, the patient is more likely to experience considerable dissatisfaction. This is demonstrated by the sharp drop in satisfaction when care investment becomes less favourable (toward the left on the x-axis). However, even delivering services of this fundamental calibre may not be sufficient to satisfy patients. Thus, the line never crosses the 0 mark, often known as “neutral satisfaction.” The implication is that more criteria are necessary to affect satisfaction. 96% of respondents in our sample were SFD or HSFD with the mannerisms of the medical personnel, the hospital staff, as well as the treatments they received. Based on these figures, it is safe to say that our survey population’s fundamental needs are being satisfied.

The model demonstrates that patients will be unsatisfied if certain quality expectations are not met and that satisfaction rises linearly as more of these expectations are met. Patients make decisions based on comparisons of these expectations to identify distinctions between rivals. In this study, 94% of the participants had access to hospitals during operating hours and were SFD or HSFD. In terms of cleanliness, 87% were SFD/HSFD, and 96% claimed the same for healthcare costs. The delighter quality curve, which only exists in the satisfaction zone, shows unexpectedly high-quality products that patients didn’t know they wanted and that, when they are, provide a pleasant surprise. In this encounter, expectations are surpassed, frequently as a result of innovation that might enhance patient expectations and give a substantial competitive advantage. Hospital data indicate that this is mostly accomplished through the patient’s perception of personalized, personalised service, supplied by compassionate and concerned doctors ^12^.

The focus of NNJS and the institutions in its network has always been patient-centred care. As an illustration, patient counselling is a service provided to particular patients who may need more help and knowledge. With 85% of patients (who used the service) reporting being SFD/HSFD, it is a service that is well-received.

Therefore, based on Kano model, patient satisfaction and extension service quality, all the relationship must be considered. This model is an effective way to explain factors affecting patient satisfaction and service quality but its limitation is it does not provide actual reason on patient’s response regarding these different factors. There are many theories that explains patient’s satisfaction in health care. Theories provide a framework on people’s perception and opinion. The two well-known theories are the healthcare quality theory (HQT) and the expectancy value theory (EVT). According to the EVT, a patient’s happiness with care is influenced by their beliefs, values, and prior expectations. The HQT, on the other hand, places a strong emphasis on the importance of the interpersonal aspects of care in promoting patient satisfaction^13^.

Regarding the variables which measured provider’s/staff relationship, there was no association, which concludes the data supports our first theory. The findings reflect respondent’s belief and background, such as respondents with greater levels of education (secondary or higher) reported feeling overall happier than respondents without any formal education. The EVT is used to support our argument that a greater education raises health literacy. When these patients attend the hospital, they have more reasonable expectations because they are better able to gather, interpret, and comprehend fundamental health information. Additionally, those with lower education levels tended to be poorer among our sample’s participants. In reality, 77% of those polled who said they couldn’t make it for more than six months on their principal source of income never attended school.

To determine the characteristics of health literacy and to evaluate its effects on patient satisfaction, preventative services, healthcare utilization, and costs, Stephanie Macleod looked at two populations, “sicker” and “healthier.”^14^. They came to the conclusion that these characteristics were negatively connected with poor health literacy, which is more prevalent in older and less educated people. Although health literacy was not a direct outcome of our study, it is conceivable that it can contribute to an understanding of the patterns in the data.

It is also conceivable that deeply ingrained cultural customs may have an impact on patient satisfaction. The conventional view of disease is that it is an outside force acting as a punishment or an external force that can only be stopped by an all-powerful force. Expectations of contemporary medicine and healthcare professionals in general are shaped by these notions. Understanding the patient population that the hospital serves is vital, especially for any misconceptions or unreasonable expectations, as a mismatch between patient expectations and the treatment obtained is linked to decreased satisfaction^15,16^. A study that’s explores the role of trust in patient satisfaction, states positive association between patient satisfaction and higher level of trust^17^.

In our sample population, 91% of those who identified as ‘DSFD’, ‘FR’, or ‘HDSFD’ overall belonged to the lowest socioeconomic class (as determined by survey criteria). Which, as was already said, may point to a lack of health literacy and possibly distorted expectations of traditional versus modern medicine. Although not every member of this group gave the same answers, it is important to note that there may be an expectation mismatch and/or a lack of trust among them. But in order to answer this, further research on these people and a comparison would be necessary. Future research on this association is important because it may shed light on the psychology of satisfaction in rural and low socioeconomic people.

## Limitation

Male patients responded to the survey more frequently than female patients (2:1) in our sample set. because we did not account for this gap in our data analysis, the satisfaction scores primarily reflect the views of men. We do not view this as a significant constraint because there was no statistical difference between the sexes. To determine the reason for this disparity and whether survey weighting would be acceptable for further studies, it is however worthwhile to review our approach used in the data collection process.

## Conclusion

According to our research, a sizable majority of patients are satisfied or extremely satisfied. All categories (staff conduct, services, and facilities) reflect this. As a result, it is safe to say that hospitals offer great perceived quality and performance. There are three theoretical curves that can be used to indicate patient satisfaction with regard to quality. Managers of hospitals should make sure that each of these tasks is carried out to its full potential. This entails fulfilling all of the patient’s fundamental requirements, including any performance requirements. The addition of delighters will also have a large effect on patient satisfaction and perceived treatment quality. Our findings suggest that background, beliefs, and individual experiences may all affect pleasure. As a result, demographic data like education level and socioeconomic position are significant factors that need to be taken into account.

## Data Availability

The dataset will be available through corresponding author.

## Acknowledgement

This research study was fully funded by Nepal Health Research Council (NHRC). We would like to thank the council for supporting us financially, tertiary eye hospital of Madhesh Province for coordination and all the respondents, who were willing to participate in this study and provide relevant information.

